# Evaluating Few-Shot Prompting for Spectrogram-Based Lung Sound Classification Using a Multimodal Language Model

**DOI:** 10.1101/2025.07.27.25332255

**Authors:** Nicholas Dietrich, Mark F. Rzepka

## Abstract

**Introduction:** Traditional deep learning models for lung sound analysis require large, labeled datasets; multimodal LLMs may offer a flexible, prompt-based alternative. This study aimed to evaluate the utility of a general-purpose multimodal LLM, GPT-4o, for lung sound classification from mel-spectrograms and assess whether a few-shot prompt approach improves performance over zero-shot prompting.

**Methods:** Using the ICBHI 2017 Respiratory Sound Database, 6898 annotated respiratory cycles were converted into mel-spectrograms. GPT-4o was prompted to classify each spectrogram in both zero-shot and few-shot settings. Few-shot prompts included labeled examples, while zero-shot prompts did not. Model outputs were evaluated against ground truth labels using performance metrics including accuracy, precision, recall, and F1-score.

**Results:** Few-shot prompting improved overall accuracy (0.363 vs. 0.320) and yielded modest gains in precision (0.316 vs. 0.283), recall (0.300 vs. 0.287), and F1-score (0.308 vs. 0.285) across labels. McNemar’s test indicated a statistically significant difference in performance between prompting strategies (p < 0.001). Model repeatability analysis demonstrated high agreement (κ = 0.76–0.88; agreement: 89–96%), indicating excellent consistency.

**Conclusion:** GPT-4o demonstrated limited but statistically significant performance gains using few-shot prompting for lung sound classification. While not yet suitable for clinical use, this prompt-based approach offers a promising, scalable strategy for medical audio analysis without task-specific training.

## Introduction

Lung sounds provide critical insights into pulmonary health and underlying disease. Traditionally, auscultation with a stethoscope has been the gold standard for assessing lung sounds [1–3]. Advancements in technology have enabled the use of electronic devices, such as digital stethoscopes and audio recorders, to capture and analyze respiratory sounds. These tools facilitate longitudinal monitoring, remote sharing, and integration into research pipelines for model development [4,5].

Recent efforts have focused on developing machine learning models for automated lung sound classification, particularly in detecting adventitious sounds such as crackles and wheezes [6,7]. A common approach involves converting audio recordings into spectrograms and applying deep learning architectures, such as convolutional neural networks (CNNs), to build binary classifiers or detection models [8–10]. While these models demonstrate promising performance, they are computationally intensive and often highly customized to specific training data, limiting their generalizability.

A potential alternative to CNNs for automated lung sound analysis is using general-purpose multimodal large language models (LLMs) [11]. Unlike traditional deep learning models, which rely on large, labeled datasets and require dedicated training for specific tasks, multimodal LLMs leverage pre-trained knowledge across diverse data formats, such as images, videos, and code. Their ability to integrate and analyze different data types may reduce the need for extensive task-specific model development. Beyond their capacity to process multimodal inputs, the performance of LLMs may be further refined through prompting strategies that optimize outputs without requiring explicit model retraining [12].

One such strategy is few-shot prompting, where a model is given a small set of labeled examples within the prompt to guide its predictions. Unlike zero-shot prompting, where no prior examples are given, few-shot learning enables models to infer patterns from limited contextual information, improving accuracy for certain complex tasks [12–14]. For example, prior research has demonstrated that few-shot prompting enhances performance in unimodal applications, such as differential diagnosis generation and text classification [15]. However, its effectiveness for multimodal tasks, particularly in the interpretation of lung sound spectrograms, remains unexplored.

To our knowledge, no prior studies have evaluated the use of a general-purpose multimodal LLM for lung sound analysis or investigated the impact of few-shot prompting in this context. This study aims to address these gaps by assessing the performance of a multimodal LLM in detecting lung sounds from mel-spectrograms and evaluating whether few-shot prompting improves classification compared to zero-shot prompting.

## Methods

### Population

This study utilized the previously validated International Conference on Biomedical and Health Informatics (ICBHI) 2017 Respiratory Sound Database [16], a publicly available dataset comprising lung sound recordings from 126 subjects across multiple clinical sites. The cohort included 46 females, 79 males, and 1 participant with missing sex data. Age distribution was as follows: 49 pediatric (<18 years), 22 adult (18–64 years), and 53 older adult (≥65 years) participants; the median age was 60.0 years (interquartile range: 4.0–70.25). Recordings were collected using four types of electronic stethoscopes and microphones. The dataset contains 920 annotated recordings segmented into 6898 respiratory cycles (inspiration and expiration), labeled as “crackles” (n=1864), “wheezes” (n=886), “both” (presence of both crackles and wheezes; n=506), or “normal” (absence of both crackles and wheezes; n=3642) based on expert consensus.

### Spectrogram Generation

Each audio file was segmented into individual respiratory cycles based on the start and end times provided in the ICBHI database [16]. Respiratory cycles were resampled to 44.1 kHz for uniformity and a custom Python pipeline was developed to generate mel-spectrograms from these cycles, applying a two-dimensional Fourier transform (2DFT) for frequency decomposition [17]. The mel-spectrograms were generated using 128 mel bands, a window size of 2048 samples, and a hop length of 512 samples, ensuring adequate time-frequency resolution. A viridis color filter was applied to enhance visual interpretability [18], and each spectrogram was structured with frequency (Hz) on the y-axis, time (s) on the x-axis, and amplitude in decibels (dB) to preserve critical acoustic features. Each spectrogram was saved in PNG format to maintain consistency in downstream analysis.

### Large Language Model Selection

GPT-4o (model GPT-4o-2024-08-06) [19] was selected as the multimodal LLM for this study due to its vision capabilities and demonstrated efficacy in prior studies involving spectrogram-based sound classification [20]. Developed by OpenAI, GPT-4o was trained on a diverse dataset that includes image, video, audio, and text modalities up to October 2023. The model was accessed via the OpenAI API, which allowed for precise control over temperature (set to 0 to minimize randomness), response length, and token constraints for deterministic outputs. The API’s batch processing capabilities enabled synchronous execution of prompts, maintaining uniformity across model queries while minimizing variability in responses.

### Prompt Engineering and Development

A standardized prompt structure was developed for both zero-shot and few-shot settings. The zero-shot prompts directly queried GPT-4o with the spectrogram and a predefined task question regarding the classification of lung sounds, ensuring that the model had no prior reference to labeled examples (Figure 1). In contrast, the few-shot prompts incorporated examples of labeled reference spectrograms, providing the model access to input-output pairs before making predictions (Figure 2). The reference spectrograms were selected from previously validated audio samples of lung sounds (crackles, wheezes, both, normal) external to the ICBHI database [21], generated with the same approach as the input spectrograms. The few-shot prompt design was based on multimodal prompting strategies from prior studies, which have been shown to improve classification accuracy when contextual examples are provided [20,22].

**Figure 1.**
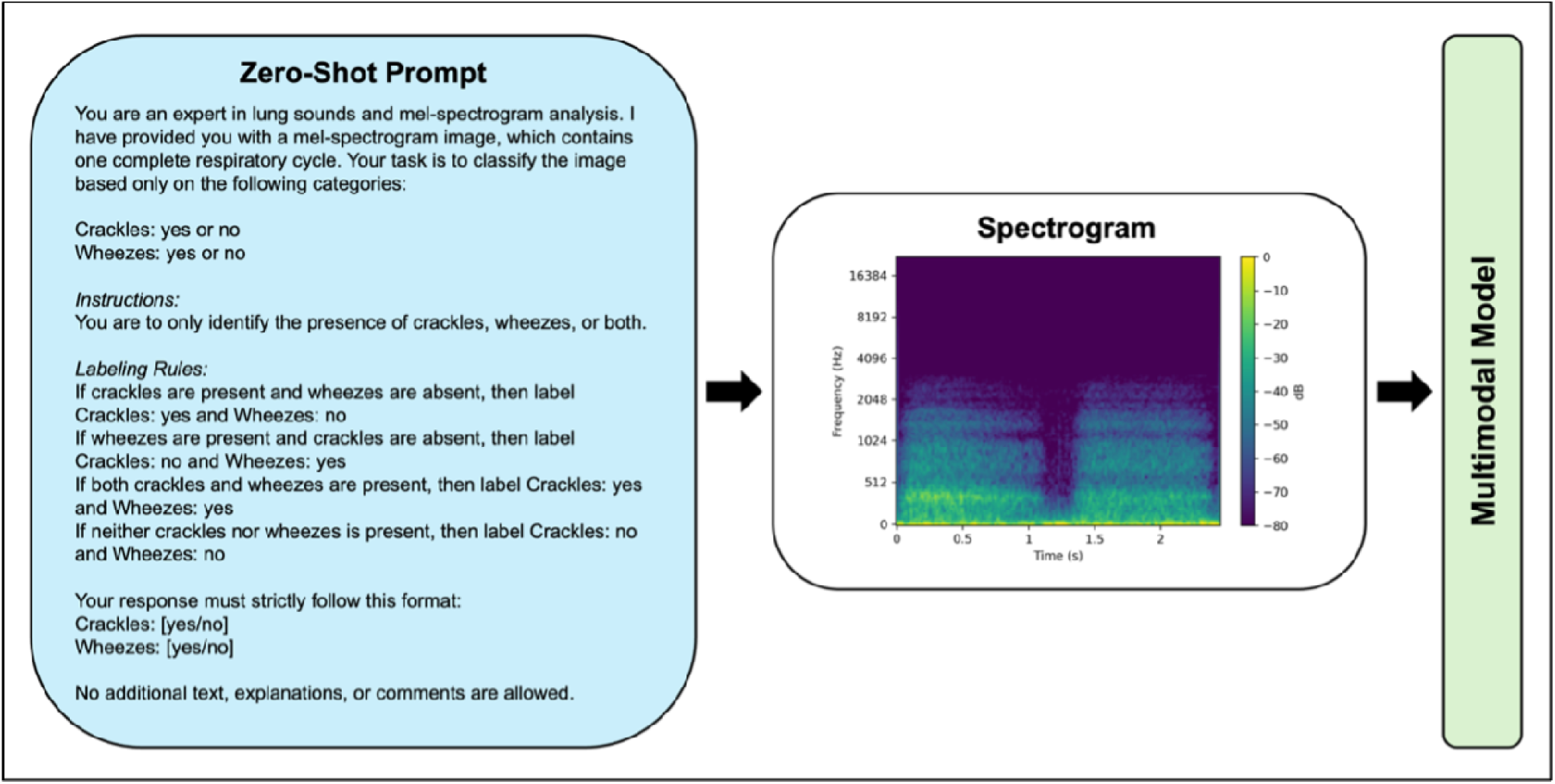
Zero-Shot Prompt Structure for Multimodal Model Lung Sound Classification.

**Figure 2.**
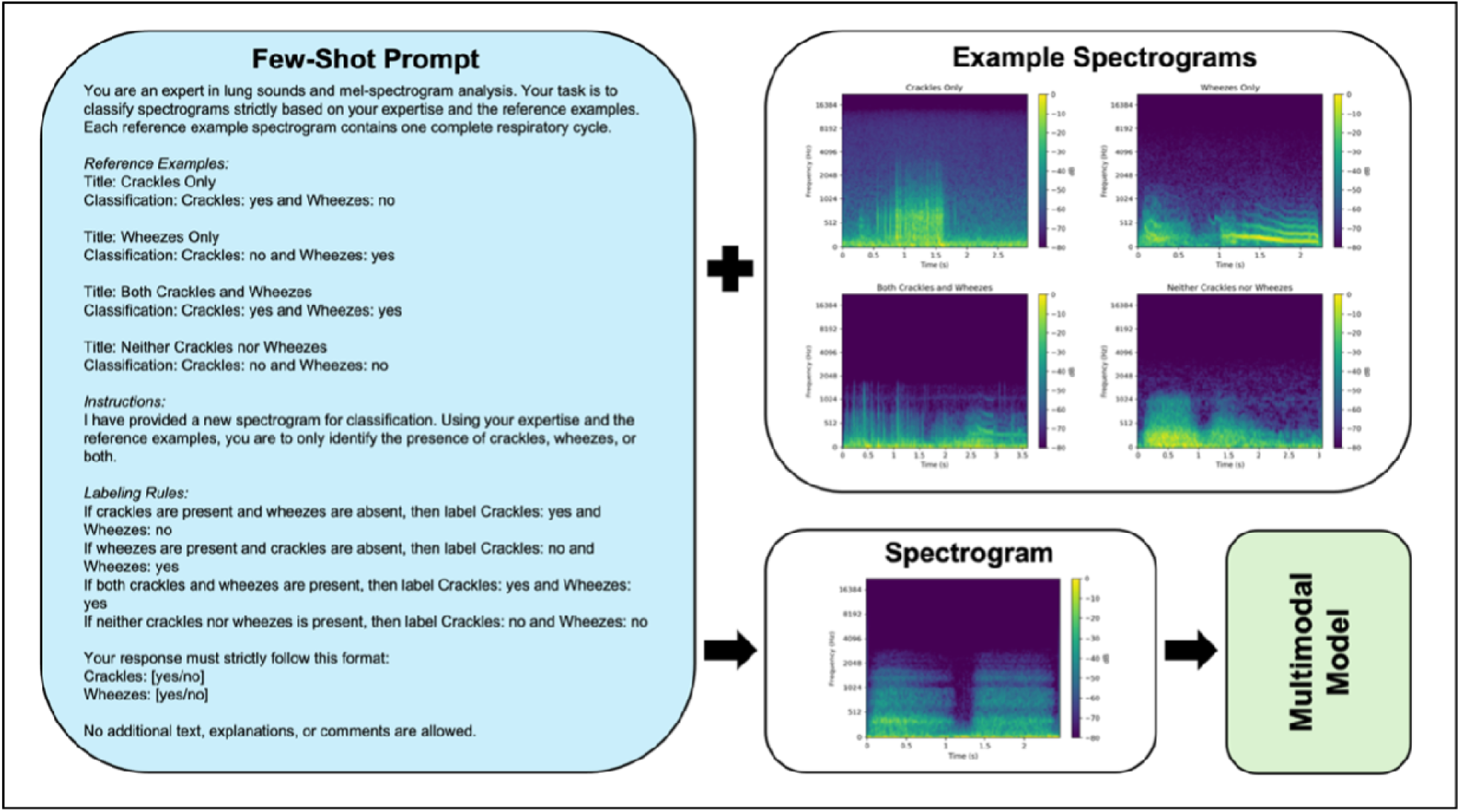
Few-Shot Prompt Structure with Labeled Reference Example Spectrograms for Multimodal Model Lung Sounds Classification.

### Data Processing Architecture

Each spectrogram was processed using the zero-shot and the few-shot prompts, with model settings and computational conditions identical across both prompting strategies. A synchronous processing pipeline was implemented to ensure that each spectrogram was sequentially processed, with each API request waiting for the previous request to complete before execution of the next. This controlled approach minimized variability and ensured that all classifications were performed under standardized conditions. The retrieved text output from GPT-4o was stored in .txt format, with post-processing implemented via a Python script to store outputs to the ICBHI annotation format [16].

### Statistical Analysis

The classification outputs from GPT-4o, using both zero-shot and few-shot techniques, were compared to the ground truth labels from the ICBHI dataset [16]. Performance metrics included accuracy, sensitivity (recall), specificity, precision (positive predictive value [PPV]), negative predictive value (NPV), and F1-score, as previously described [8,23]. These metrics were calculated both overall and on a per-label basis. To evaluate differences in the overall classification performance between the zero-shot and few-shot settings, McNemar’s test was applied. Statistical significance was set at a two-sided p-value of 0.05.

For the per-label analysis, two analytical approaches were employed [16,23]. First, in the multiclass classification approach, the model was treated as a four-label classifier with mutually exclusive categories: only crackles, only wheezes, both, and normal. Second, in the binary classification approach, the model was treated as two independent binary classifiers, one for crackles (present/absent) and one for wheezes (present/absent), allowing for separate identification of each sound even when both were present in a single respiratory cycle. We also conducted a subgroup analysis for the “both” and “normal” ground truth labels to compare model performance on detecting crackles versus wheezes within these categories. This included calculating the frequency of each label prediction and evaluating differences between zero- and few-shot prompting using McNemar’s test for paired binary outcomes.

To assess model output repeatability, 10% of the dataset was randomly selected and re-evaluated using the same zero- and few-shot prompts under identical model conditions. Classification labels for crackles and wheezes were compared between the initial and repeated outputs. For each condition, both Cohen’s kappa and percentage agreement were calculated to quantify the consistency between runs.

## Results

### Overall Classification Performance

Overall classification accuracy was 0.320 (95% confidence interval [CI]: 0.309–0.331) for zero-shot prompting and 0.363 (95% CI: 0.352–0.374) for few-shot prompting. For the zero-shot prompting, the model achieved a true positive (TP) count of 935, true negative (TN) of 1271, false positive (FP) of 2371, and false negative (FN) of 2321. Corresponding performance metrics were: precision 0.283 (95% CI: 0.267–0.298), recall 0.287 (95% CI: 0.272–0.303), F1-score 0.285, specificity 0.349 (95% CI: 0.334–0.364), and NPV 0.354 (95% CI: 0.338–0.369).

Using few-shot prompting, the model produced counts of 976 TP, 1529 TN, 2113 FP, and 2280 FN. Performance improved across most metrics: precision was 0.316 (95% CI: 0.300–0.332), recall 0.300 (95% CI: 0.284–0.315), F1-score 0.308, specificity 0.420 (95% CI: 0.404–0.436), and NPV 0.401 (95% CI: 0.386–0.417). McNemar’s test revealed a statistically significant difference in performance (χ^2^ = 86.47, p < 0.001), indicating that the few-shot model significantly outperformed the zero-shot model.

The repeatability analysis demonstrated Cohen’s kappa values of 0.88 and 0.78 for zero-shot crackles and wheezes, respectively, and 0.83 and 0.76 for few-shot crackles and wheezes, indicating substantial agreement across model runs. Agreement percentages were high for all comparisons, 95.5% for zero-shot crackles, 89.3% for zero-shot wheezes, 92.6% for few-shot crackles, and 90.6% for few-shot wheezes. No statistically significant differences were observed in overall classification performance between the initial and repeated runs (p>0.05), supporting the consistency and reliability of model predictions across repeated evaluations.

### Per-Label Classification Performance

In the multiclass analysis, accuracy for the “only crackles” label was 0.603 (95% CI: 0.592– 0.615) for zero-shot prompting and 0.582 (CI: 0.571–0.594) for few-shot prompting. For “only wheezes,” accuracy improved from 0.634 (95% CI: 0.623–0.645) in the zero-shot model to 0.737 (95% CI: 0.726–0.747) in the few-shot model. The “both” category showed high accuracy for both models, with 0.924 (95% CI: 0.918–0.931) for zero-shot and 0.915 (95% CI: 0.909–0.922) for few-shot prompting. Accuracy for the “normal” label was 0.478 (95% CI: 0.466–0.490) for zero-shot and 0.492 (95% CI: 0.480–0.504) for few-shot prompting.

Using the binary classification analysis, accuracy for detecting crackles was 0.563 (95% CI: 0.552–0.575) in the zero-shot model and 0.545 (95% CI: 0.533–0.557) in the few-shot model. For wheezes, accuracy improved from 0.628 (95% CI: 0.617–0.640) in the zero-shot setting to 0.710 (95% CI: 0.700–0.721) with few-shot prompting. The remaining per-label performance metrics are reported in Table 1 for the four classes and Table 2 for the two binary classes.

**Table 1.**
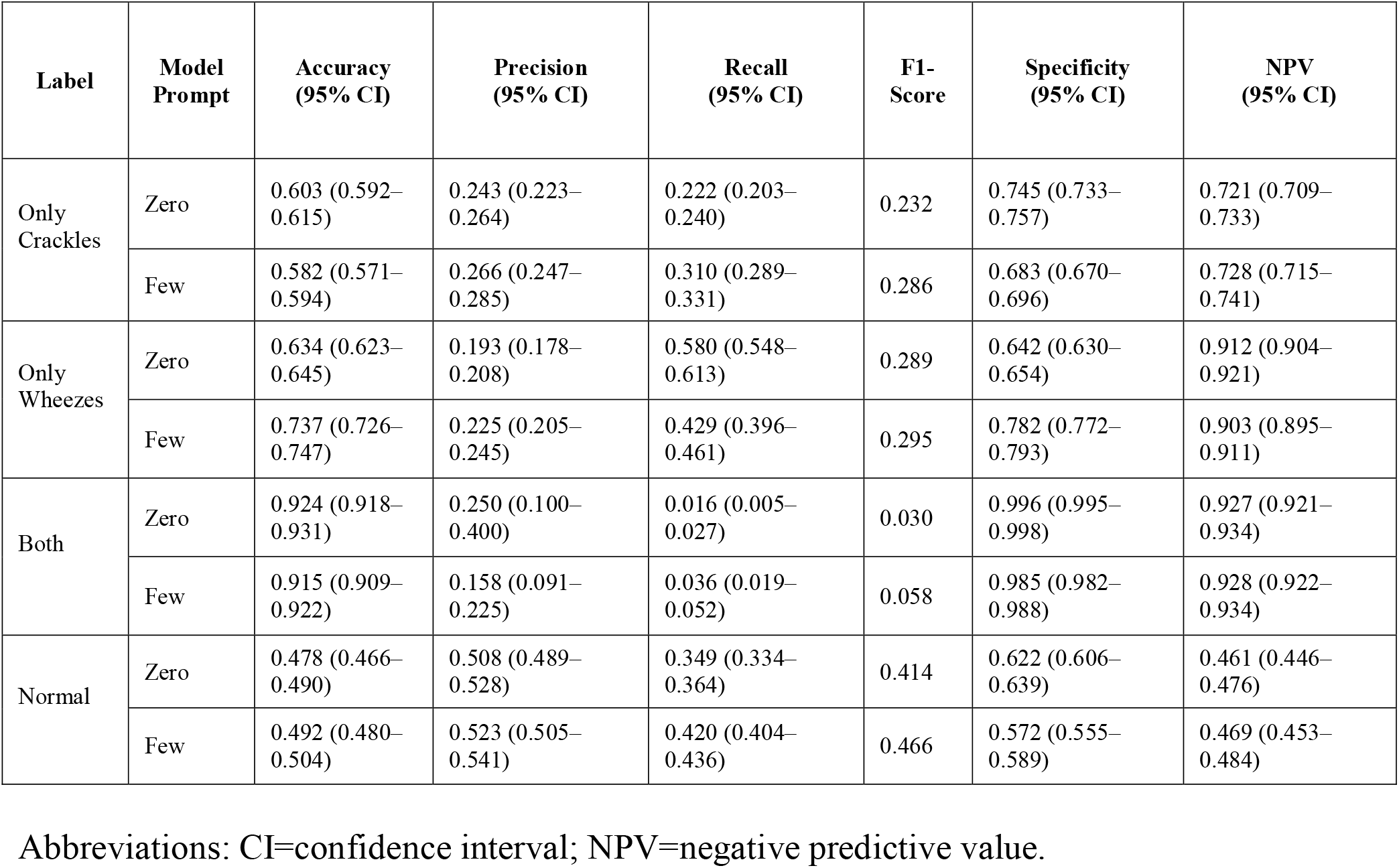
Multiclass Performance Metrics for Each Lung Sound Label (Only Crackles, Only Wheezes, Both, Normal) Using Zero-Shot and Few-Shot Prompting.

| Label | Model Prompt | Accuracy (95% CI) | Precision (95% CI) | Recall (95% CI) | F1-Score | Specificity (95% CI) | NPV (95% CI) |
| --- | --- | --- | --- | --- | --- | --- | --- |
| Only Crackles | Zero | 0.603 (0.592–0.615) | 0.243 (0.223–0.264) | 0.222 (0.203–0.240) | 0.232 | 0.745 (0.733–0.757) | 0.721 (0.709–0.733) |
|  | Few | 0.582 (0.571–0.594) | 0.266 (0.247–0.285) | 0.310 (0.289–0.331) | 0.286 | 0.683 (0.670–0.696) | 0.728 (0.715–0.741) |
| Only Wheezes | Zero | 0.634 (0.623–0.645) | 0.193 (0.178–0.208) | 0.580 (0.548–0.613) | 0.289 | 0.642 (0.630–0.654) | 0.912 (0.904–0.921) |
|  | Few | 0.737 (0.726–0.747) | 0.225 (0.205–0.245) | 0.429 (0.396–0.461) | 0.295 | 0.782 (0.772–0.793) | 0.903 (0.895–0.911) |
| Both | Zero | 0.924 (0.918–0.931) | 0.250 (0.100–0.400) | 0.016 (0.005–0.027) | 0.030 | 0.996 (0.995–0.998) | 0.927 (0.921–0.934) |
|  | Few | 0.915 (0.909–0.922) | 0.158 (0.091–0.225) | 0.036 (0.019–0.052) | 0.058 | 0.985 (0.982–0.988) | 0.928 (0.922–0.934) |
| Normal | Zero | 0.478 (0.466–0.490) | 0.508 (0.489–0.528) | 0.349 (0.334–0.364) | 0.414 | 0.622 (0.606–0.639) | 0.461 (0.446–0.476) |
|  | Few | 0.492 (0.480–0.504) | 0.523 (0.505–0.541) | 0.420 (0.404–0.436) | 0.466 | 0.572 (0.555–0.589) | 0.469 (0.453–0.484) |

Abbreviations: CI=confidence interval; NPV=negative predictive value.

**Table 2.**
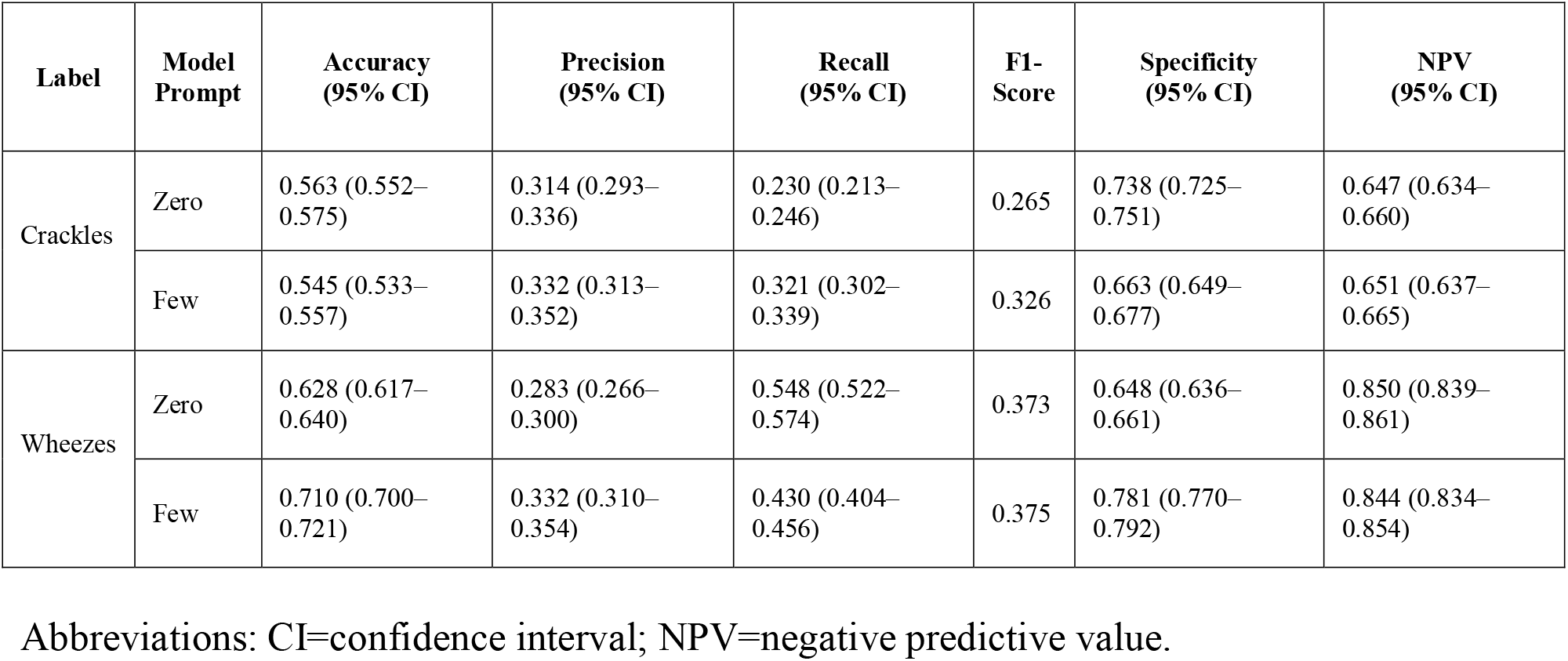
Binary Classification Performance Metrics for Crackles and Wheezes Using Zero-Shot and Few-Shot Prompting.

On sub-analysis of the classification performance for the “both” label (where both crackles and wheezes were present in the ground truth), the model identified crackles in 126 zero-shot and 170 few-shot cases, and wheezes in 244 zero-shot and 186 few-shot cases. Only 8 instances had both crackles and wheezes correctly predicted in the zero-shot setting, compared to 18 in the few-shot setting. McNemar’s test revealed statistically significant differences in performance between zero- and few-shot models for both crackles (χ^2^ = 27.191, p < 0.001) and wheezes (χ^2^ = 38.679, p < 0.001).

For the “normal” label (where both crackles and wheezes were absent), the model correctly predicted absence of crackles in 2,643 zero-shot and 2,383 few-shot cases, and absence of wheezes in 2,256 and 2,737 cases, respectively. Both crackles and wheezes were jointly absent in 1,271 zero-shot and 1,529 few-shot cases. Again, McNemar’s test showed significant differences between zero- and few-shot prompting for both crackles (χ^2^ = 140.337, p < 0.001) and wheezes (χ^2^ = 344.395, p < 0.001).

## Discussion

To our knowledge, this study is the first to assess the classification of lung sound spectrograms using a general-purpose multimodal LLM and to evaluate the performance of few-shot prompting compared to zero-shot prompting in a lung auscultation context. Our findings demonstrate reliably that, while overall model performance was low, few-shot prompting significantly outperformed zero-shot prompting across several metrics including accuracy, specificity, and F1-score for the majority of lung sound identification tasks. Consistent with findings suggesting that text-based few-shot prompting can improve diagnostic abilities of multimodal LLMs, our results importantly find that this effect can also be captured for image-based few-shot prompting [22]. Consequently, our study supports the claim that multimodal LLMs can learn task-relevant visual features through contextual exposure, even without task-specific training [24].

Unsurprisingly, the overall performance of both prompting strategies fell below clinically acceptable thresholds, with overall accuracy for zero-shot and few-shot settings at 32.0% and 36.3%, respectively [8]. These values are also notably lower than those reported in prior work using task-specific deep learning models for lung sound classification for ICBHI dataset [16]. For example, CNNs trained on spectrograms from the same ICBHI dataset have achieved accuracy scores ranging from around 50% to 90% depending on preprocessing, model architecture, and class balancing techniques [8,23]. This illustrates a substantial performance gap between specialized CNN models and general-purpose multimodal models at their current stage of development.

Regarding individual lung sound labels, performance was more pronounced in the classification of wheezes than crackles. This may be due to the more continuous and tonal nature of wheezes, which manifest in spectrograms as sustained horizontal patterns, features that may be more visually distinctive to a multimodal LLM. Crackles, on the other hand, are brief, discontinuous events whose sporadic nature may be less readily interpretable by a vision model without prior domain training [25]. This visual distinction could explain why recall for crackles remained relatively low, even in the few-shot condition (0.321).

An area of poor individual labeling performance was in identifying lung cycles containing both crackles and wheezes, underscoring a key challenge in multimodal LLM classification: integrating compound acoustic features. The multiclass results showed that both prompting strategies had difficulty recognizing this dual-label class, with recall below 4% in both cases and F1-scores near zero (however, few-shot prompting impressively more than doubled recall for this classification). These results suggest that when features overlap or co-occur with considerable variety, GPT-4o may struggle to disentangle individual features without task-specific tuning [26]. This is in contrast with the “normal” label, where the model showed reasonable performance identifying spectrograms lacking adventitious sounds. A likely explanation for this effect is the relative visual uniformity of normal breath sound spectrograms, without the unpredictable occurrence of wheezes or crackles within a respiratory cycle [25].

Collectively, our results highlight both the promise and the limitations of using general-purpose LLMs like GPT-4o for medical image-based classification tasks. While performance is inferior to task-specific deep learning approaches, the model’s responsiveness to few-shot prompting is reasonable considering GPT-4o’s lack of domain-specific training. Interestingly, the improvement gained by few-shot prompting on diagnostic accuracy compared with zero-shot prompting for multimodal LLMs remains relatively greater for text-based few-shot prompts rather than the image-based few-shot prompting performed in this study [22]. To improve performance of lung sound spectrogram detection by multimodal LLMs, future studies could further optimize few-shot prompt strategies. This could include the selection of more visually distinctive exemplars, leveraging semantic embeddings for retrieval-augmented prompting, and refining language instructions for multimodal input alignment [27–29]. These efforts may be particularly relevant in low-resource settings or in research environments where labeled data is scarce or difficult to acquire [30]. Future research should also explore the integration of these tools into clinical settings, with a focus on evaluating performance across diverse patient populations, including real-world, low-quality recordings and additional adventitious lung sounds such as rhonchi and stridor [31,32].

This study was not without its limitations. The spectrograms generated from the ICBHI 2017 database for the purpose of standardizing visual representations of lung sounds, may not have captured all acoustic nuances, especially in the presence of background noise [16]. However, these unideal features reflect real-world variability and support external generalizability. While the zero- and few-shot prompts followed previously validated approaches and were applied consistently across all cases, the specific prompt design may influence model output, and alternative prompt formulations could yield different results [28]. Lastly, we only tested GPT-4o, which, although a widely used and well-studied foundational model, may not reflect the performance of smaller or domain-specific models trained on relevant medical data [33].

## Conclusion

Our study demonstrates that general-purpose multimodal LLMs can be utilized for lung sound classification from mel-spectrograms inputs. Few-shot prompting significantly improved performance compared to zero-shot prompting across multiple metrics. While still inferior to task-specific deep learning models, this prompt-based approach demonstrates potential as a flexible and scalable alternative. Future research should investigate techniques to improve prompt performance and validate findings across diverse clinical datasets in order to enhance accuracy and, ultimately, clinical utility.

## Data Availability

All data produced in the present study are available upon reasonable request to the authors.

